# Gestational-age-dependent development of the neonatal metabolome

**DOI:** 10.1101/2020.03.27.20045534

**Authors:** Madeleine Ernst, Simon Rogers, Ulrik Lausten-Thomsen, Anders Björkbom, Susan Svane Laursen, Julie Courraud, Anders Børglum, Merete Nordentoft, Thomas Werge, Preben Bo Mortensen, David M Hougaard, Arieh S.Cohen

**Author notes:** **Corresponding authors:** Madeleine Ernst and Arieh S. Cohen, Section for Clinical Mass Spectrometry, Department of Congenital Disorders, Danish Center for Neonatal Screening, Statens Serum Institut, 5 Artillerivej, 2300 Copenhagen, Denmark, [ and ].

## Abstract

**Background:** Prematurity is a severe pathophysiological condition associated with increased morbidity and mortality; however, little is known about the gestational-age-dependent development of the neonatal metabolome.

**Methods:** Using an untargeted liquid chromatography tandem mass spectrometry (LC-MS/MS) metabolomics protocol we measured over 6000 metabolites in 148 neonatal heel prick dried blood spots retrieved from the Danish Neonatal Screening Biobank. Using a combination of state-of-the-art metabolome mining tools, including mass spectral molecular networking (GNPS), unsupervised substructure discovery (MS2LDA) and *in silico* structure annotation, we retrieved chemical structural information at a broad level for over 4000 (60%) metabolites and assessed their relation to gestational age.

**Results:** A total of 744 (∼12%) metabolites were significantly correlated with gestational age (false-discovery-rate-adjusted *P* < 0.05), whereas 93 metabolites were strongly predictive of gestational age, explaining on average 37% of the variance. Using a custom algorithm based on hypergeometric testing we identified 17 molecular families (230 metabolites) overrepresented with metabolites correlating with gestational age (*P* < 0.01). Metabolites significantly related to gestational age included bile acids, carnitines, polyamines, amino-acid-derived compounds, nucleotides, dipeptides as well as treatment-related metabolites such as antibiotics and caffeine.

**Conclusions:** Carnitines, bile acids, as well as amino acid-derived compounds are known to be affected by the gut microbiota, whereas polyamines such as spermine and spermidine may play an important role in regulating (epithelial) cell growth. Our findings reveal for the first time the gestational-age-dependent development of the neonatal blood metabolome and suggest that gut microbial and gestational-age-dependent metabolic maturation may be monitored during newborn screening.

## Introduction

Prematurity is a complex and challenging pathophysiological condition associated with increased morbidity and mortality (1,2). Numerous early- and late onset disorders are associated with preterm birth, including psychiatric disorders (9). The risk of complications is inversely correlated to the gestational age at birth but with large variations within age-groups. It is likely that the individual maturation more accurately represents the complication risk. Metabolomic analyses could allow for an individual assessment of multi-organ function and maturity and may thus contribute to improved understanding of pathophysiological mechanisms behind the development of prematurity associated disorders. However, currently there is limited knowledge on the metabolomic profile of preterm neonates (3–5) and on how to assess metabolic maturity.

Growing evidence suggests a strong link between early-life microbiota and disease (6) and short-as well as long-term complications associated with preterm birth such as necrotizing enterocolitis, diabetes, cardiovascular disease, neurodevelopmental disorders and neuropsychiatric disorders have been related to the underdevelopment of the gut microbiota, gastrointestinal tract and immune system (7–13). Although the exact timing of the establishment of the intestinal microbiome in human life remains unknown (14), it is generally agreed upon that the gut microbial colonization starts at birth at the latest and undergoes shifts in composition and structure as the host matures over time (15–19).

Recent studies have shown that marker metabolites of microbial metabolism are readily detectable in human blood and that the human blood metabolome may predict gut bacterial α-diversity (20). However, gut microbiome-derived metabolites that become available to the preterm infant and may impact gut maturation and overall host metabolism and health remain unknown.

Monitoring gut microbial health and metabolic maturation during newborn screening could offer a powerful tool for the early detection and possible early intervention in disease progression through probiotics, diet or microbial transplants. Dried blood spots (DBS) routinely collected for newborn screening are minimally invasive and metabolomics approaches enable the simultaneous measurement of thousands of metabolites thus offering unique insights into metabolic underpinnings of complex pathophysiological conditions (21,22).

Here we hypothesize that gestational-age dependent microbial and fetal maturation impacting overall host metabolism and health may be monitored during newborn screening. Using dried blood spots retrieved from the Danish Neonatal Screening Biobank in combination with recently developed computational metabolomics tools including mass spectral molecular networking, unsupervised substructure discovery and *in silico* structure annotation (23–26), we assess the gestational age-dependent development of the blood metabolome of very preterm to term neonates.

## Methods

### Study Cohort

Residual extracts from the Danish newborn screening program during the period from March to June 2016 were selected for this study. A total of 148 newborns (62 girls) were sampled at 48-72h of age, with gestational ages from 28 to 42 weeks (n = 8-10 each) (Supplementary Table 1). The study was conducted in accordance with the Declaration of Helsinki and the protocol complies with the Danish Ethical Committee law by not being a health research project (§2,1) but a method development study not requiring an ethical approval (27).

### Metabolomic Profiling

All samples (including blank and pooled quality control samples) were submitted to untargeted metabolomic profiling using liquid chromatography tandem mass spectrometry (LC-MS/MS) at Statens Serum Institut, Copenhagen, Denmark between July 6, 2016 and July 14, 2016. Raw data files were preprocessed using MZmine (version 2.40.1) (28). A detailed description of LC-MS/MS as well as preprocessing parameters can be found in the Supplementary Methods.

### Statistical Analyses

Overall variation in the metabolome related to gestational age was assessed using a principal coordinates analysis (PCoA) plot with the Bray-Curtis dissimilarity. A Permutational Multivariate Analysis of Variance (PERMANOVA) (29) model was fitted to the Bray-Curtis distance matrix to assess the variation in the metabolome explained by gestational age. To predict gestational age from the neonatal metabolome, we used a tenfold cross-validation (CV) implementation of the least absolute shrinkage and selection operator method (LASSO), including comparing its performance to a Ridge regression such as described in Wilmanski and co-workers (20). Metabolite richness and α-diversity was assessed using the mean number of metabolites and Shannon Index, respectively, measured per sample and stratified by categories of prematurity and gestational age. Subsequently, a Kruskal-Wallis test was used to compare mean metabolite richness and α-diversity per prematurity category, whereas correlation between metabolite richness and gestational age was evaluated using Kendall’s Tau (30).

Univariate correlation at the individual metabolite level was assessed using Kendall’s Tau and P values were adjusted for multiple hypothesis testing using the false discovery rate (FDR) method (31). Hypergeometric testing was used to identify chemically structurally related molecular families overrepresented with metabolites that had a statistically significant correlation with gestational age. Additional information is provided in the Supplementary Methods. All statistical analyses were performed in R 3.6.1 (32) or Python 3.7 (33). All Jupyter notebooks used for statistical analysis are publicly available at: https://github.com/madeleineernst/Prematurity_SupplementaryMaterial.

### Metabolite Identification

Aggregated MZmine preprocessed MS/MS fragmentation spectra were submitted to feature-based mass spectral molecular networking through the Global Natural Products Social Molecular Networking Platform (GNPS) (24,34,35) and searched against all GNPS spectral libraries. To further enhance chemical structural information within the network, substructure information was incorporated using the GNPS MS2LDA workflow (https://ccms-ucsd.github.io/GNPSDocumentation/ms2lda/) (23). Information from *in silico* structure annotations from Network Annotation Propagation (25) and Dereplicator (36) were incorporated using the GNPS MolNetEnhancer workflow (https://ccms-ucsd.github.io/GNPSDocumentation/molnetenhancer/) (26) with chemical class annotations retrieved from the ClassyFire chemical ontology (37). A detailed description of all workflow parameters can be found in the Supplementary Methods. Mass spectral molecular network data, data from MS2LDA unsupervised substructure discovery and *in silico* structure annotation are available upon request.

## Results

A total of 6053 metabolites (mass spectral features with unique MS/MS fragmentation patterns) were measured. Using a combination of metabolome mining tools, including mass spectral molecular networking (GNPS), unsupervised substructure discovery (MS2LDA) and *in silico* annotation through the MolNetEnhancer workflow (26), chemical structural information at the chemical class level, corresponding to a level 3 metabolite identification according to the Metabolomics Standard Initiative’s reporting standards (38) could be retrieved for nearly 60% (4176) of the detected metabolites (Supplementary Figure 1).

**Figure 1.**
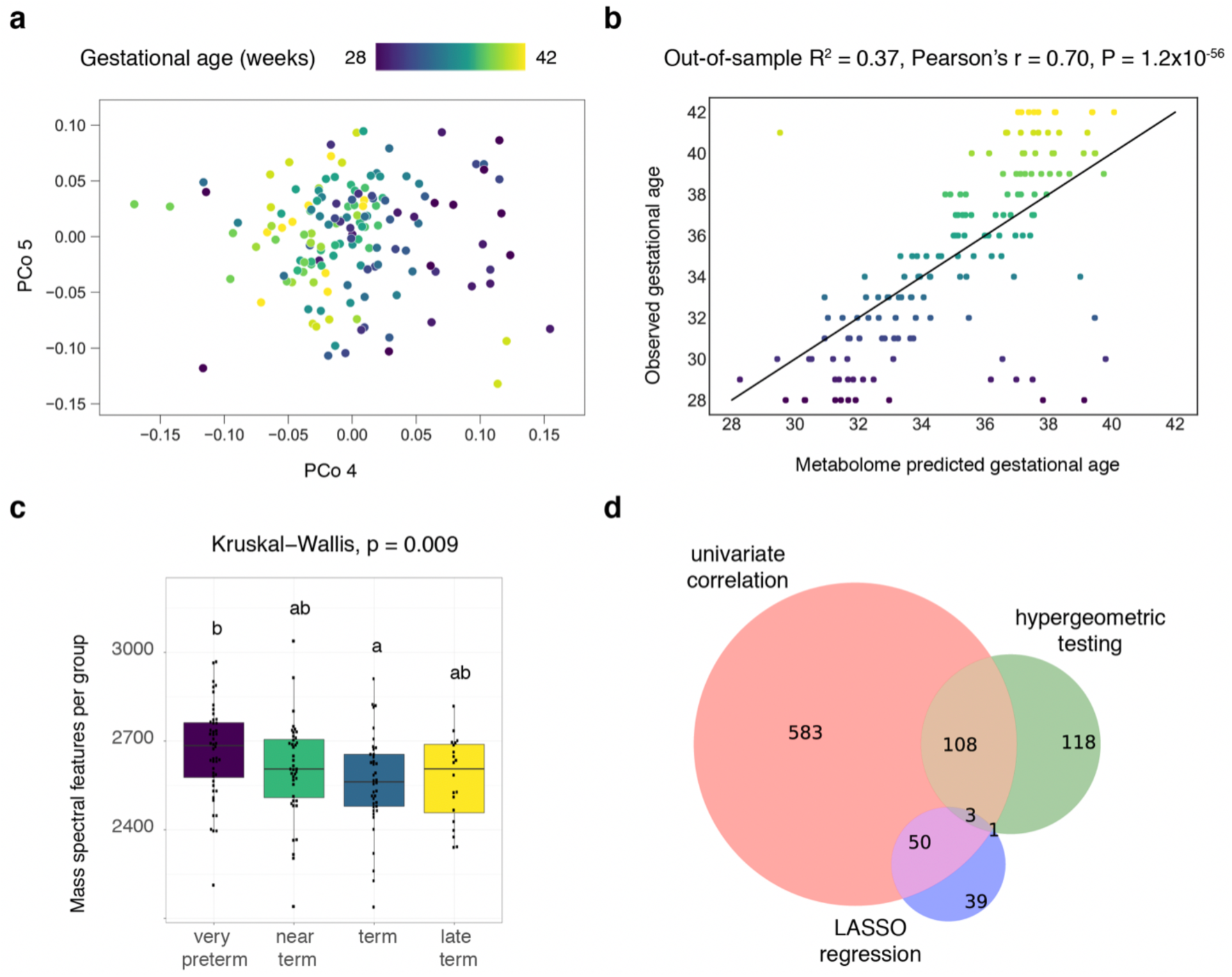
**a**. Principal coordinates analysis using the Bray-Curtis distance metric. 3.7% of the variation in the data is explained by gestational age (PERMANOVA, P < 0.05, Adonis R^2^ = 0.037). **b**. Metabolome-predicted gestational age versus observed, ultrasound-guided gestational age. Mean R^2^ across ten cross-validations, Pearson’s correlation coefficient and P-value are shown. **c**. Box plots for number of mass spectral features stratified across different prematurity categories. Significant differences were found across mean number of mass spectral features per prematurity category (very preterm: 28-23 weeks; near term: 33-36 weeks; term: 37-40 weeks; late term: 41-42 weeks) (Kruskal-Wallis, P < 0.05). **d**. Venn diagram illustrating overlapping metabolites significantly associated with gestational age by univariate correlation analysis, hypergeometric testing at the molecular family level and LASSO regression.

Principal coordinates analysis (PCoA) and permutational analysis of variance demonstrated that 3.7% of the variation in the metabolomics data could be explained by gestational age (PERMANOVA, P < 0.05, Adonis R^2^ = 0.037) with strongest separation observed along principal coordinate (PCo) 4 (Figure 1a).

The LASSO model suggested that a total of 93 metabolites were strongly predictive of gestational age, explaining an average of 37% of the variance (mean out-of-sample R^2^ = 0.37, Pearson’s r = 0.70, P = 1.2 × 10^−22^ for metabolome-predicted gestational age versus observed, ultrasound-guided gestational age) (Figure 1b, Supplementary Results). Out of these 93 metabolites, six could be matched to GNPS library spectra, including pyroglutamylvaline, 1-hexadecylpyridinium, serine, ophthalmic acid, N1-acetylspermine and spermidine. Four unannotated metabolites were selected in all tenfold cross-validation (CV) models and were the most influential in predicting gestational age, whereas 93 metabolites were retained by at least one model. No chemical structural information could be retrieved for the four metabolites selected in all tenfold cross-validation models. However, by comparing mass spectral fragmentation spectra to data in the public domain (39), we found that one of the four metabolites was previously found in human sputum samples of patients with cystic fibrosis undergoing antibiotic treatment, fecal samples of children and the surface of a tomato plant. This suggests that the unknown structure is likely of (anti)-microbial nature.

Mean metabolite richness was found to vary significantly with different categories of prematurity (Kruskal-Wallis, P < 0.05), with higher numbers of metabolites observed for very preterm versus term neonates (Figure 1c). Furthermore, mean metabolite richness was found to correlate significantly with gestational age (Kendall’s Tau = -0.2, P < 0.05) with more metabolites observed in neonates born at 28 weeks of gestation (Supplementary Figure 2). Within sample metabolite diversity (Shannon Index) on the other hand was not found to vary significantly with different categories of prematurity (Kruskal-Wallis, P = 0.15) (Supplementary Figure 3).

**Figure 2.**
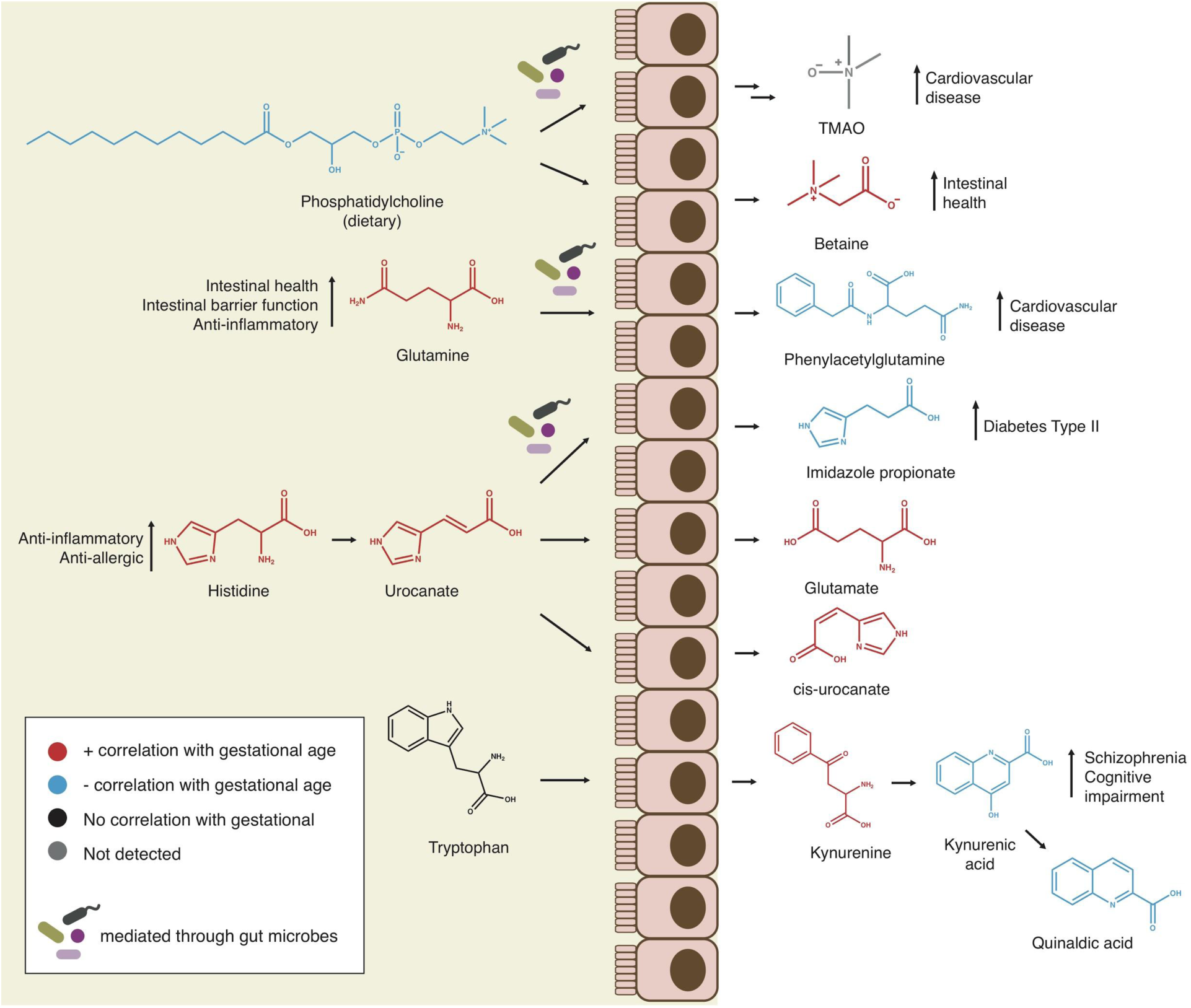
Schematic representation of amino acid and phospholipid catabolism pathways in the gut with metabolites significantly correlating with gestational age highlighted. Molecules highlighted in red are positively correlated with gestational age, whereas molecules highlighted in blue are negatively correlated with gestational age (Kendall’s Tau, false-discovery-rate-adjusted P < 0.05). Molecules more abundant in preterm neonates (negative correlation with gestational age), have previously been associated with health complications related to prematurity, such as cardiovascular diseases, diabetes and cognitive impairment, and in three out of four cases may be mediated through gut microbiota.

**Figure 3.**
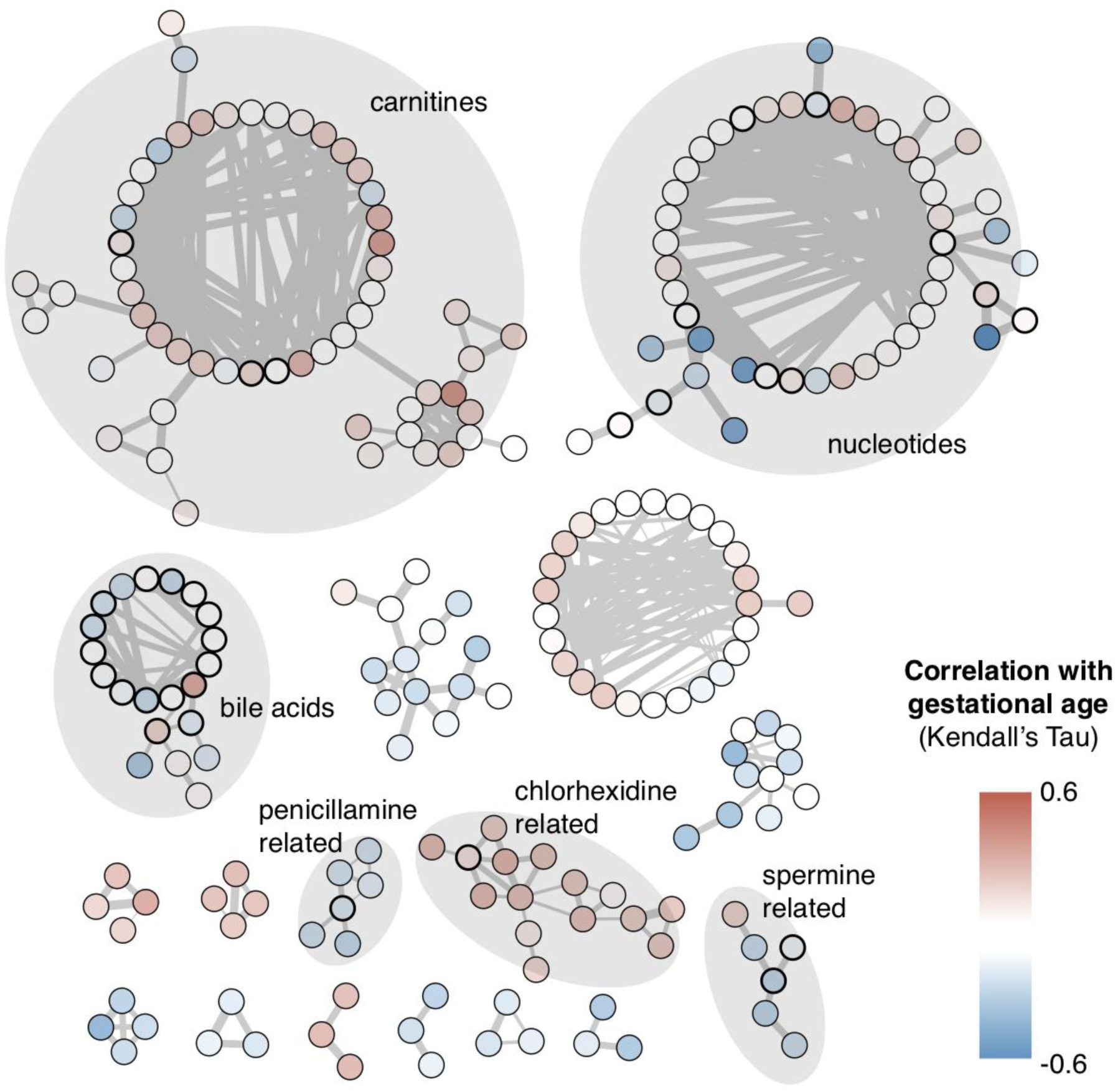
Molecular families significantly overrepresented in metabolites correlating with gestational age (P < 0.01) identified through hypergeometric testing. Node colors represent correlation with gestational age (Kendall’s Tau). Metabolites for which chemical structural annotation could be retrieved are indicated with grey shadowing. The thickness of the lines connecting the nodes represents tandem mass spectral similarity, implying high chemical structural similarity. Nodes with bold black borders indicate GNPS spectral library hits. Carnitines, bile acids, nucleotides and spermine were previously reported to be implicated in the maturation of the gastrointestinal tract or being affected by the gut microbiome. Penicillamine could be reflective of antibiotics use and chlorhexidine, a common skin disinfectant reflective of different sampling strategies across term and preterm neonates.

In the univariate analyses, 744 metabolites (∼12%) were found to significantly correlate with gestational age (FDR-adjusted P < 0.05, Supplementary Figure 4). Out of these 744 metabolites, 100 could be chemically structurally annotated through GNPS library matching, manual or *in silico* annotation propagation throughout the mass spectral molecular network, including amino acids, carbohydrates (sugars), dipeptides, lipids (carnitines, bile acids and phospholipid catabolites), nucleotides, polyamines (N1-acetylspermine, spermine, spermidine and structural analogues) and xenobiotics (caffeine, antibiotic-derived metabolites including penicillamine disulfide and cefuroxime as well as compounds related to chlorhexidine, a common disinfectant) (Supplementary Data 3). Out of the ten amino acids significantly correlating with gestational age, nine were found to possibly be related through the histidine, tryptophan or phenylalanine and tyrosine metabolic pathways (40–43), respectively (Supplementary Data 3, Figure 2). Six amino acids or catabolites were found to correlate positively with gestational age (*cis*-urocanate, glutamine, glutamic acid, histidine, kynurenine, serine), whereas four were found to correlate negatively with gestational age (imidazole propionate, kynurenic acid, phenylacetylglutamine, quinaldic acid). Betaine and phosphatidylcholine were found to be possibly biosynthetically related through the phospholipid catabolism, with betaine increasing and phosphatidylcholine decreasing with gestational age. All peptides, most carnitine and chlorhexidine structural analogues were found to increase with gestational age, whereas antibiotics, caffeine, bile acids (except for hyocholic acid), most nucleotide structural analogues, polyamines, and sugars were found to decrease with gestational age.

A total of 17 molecular families, comprising 230 metabolites were found to be significantly overrepresented (P < 0.01) with metabolites correlating with gestational age (non-FDR-adjusted P < 0.01) by hypergeometric testing (Figure 3). Chemical structural annotation could be retrieved for 6 molecular families (152 metabolites) and revealed that carnitine families mostly correlated positively with gestational age, whereas nucleotide, bile acid and spermine-related families correlated negatively with gestational age (Supplementary Figures 5 to 9). A family of structural analogues of penicillamine, likely a degradation product of penicillin, was found to correlate negatively with gestational age while structural analogues of chlorhexidine, a common disinfectant, were found to correlate positively with gestational age. Three metabolites (two nucleotides and spermidine) were identified by all three statistical approaches to be significantly associated with gestational age (Figure 1d).

## Discussion

There is currently limited knowledge on the metabolomic status of preterm neonates and a deeper understanding hereof will help further elucidate the complex pathophysiology of prematurity.

In this methodological study, the new tools deployed on untargeted metabolomics data from newborn dried blood spots were demonstrated to have great potential to address future research questions. While a number of studies have assessed variation of a few selected metabolites with gestational age (e.g. (44,45)), this is the first study assessing metabolome-wide changes related to prematurity in dried blood spots from newborn screening (4).

We found that the metabolome of 2-3-day-old neonates is highly reflective of gestational age. Using statistical modeling, univariate correlation analysis at the individual metabolite level and hypergeometric testing at the molecular family level we found that 93, 744 and 17 molecular families (constituting 230 metabolites), respectively, were significantly associated with gestational age.

On average only 2-5% of metabolites can be chemically structurally annotated in untargeted LC-MS/MS based metabolomics studies (46). Using a combination of different computational metabolomics tools, we were able to retrieve chemical structural information at a broad level for nearly 60% of the data collected, thus representing a major advance in biochemical interpretation. Furthermore, evaluating changes at the level of chemically structurally related molecular families (rather than individual metabolites) through hypergeometric testing is a novel approach, which allows to understand metabolic changes of groups of metabolites changing consistently but modestly across samples. This approach increases the chance for novel discoveries and a potentially better understanding of pathobiological processes as these metabolites would be missed in univariate approaches.

We identified amino-acid-derived metabolites, dipeptides, polyamines, nucleotides, lipids (bile acids, phosphatidylcholine-derived compounds), sugars, and treatment-related compounds such as penicillamine disulfide, cefuroxime and caffeine as significantly correlating with gestational age. Similarly, LASSO regression identified dipeptides, polyamines, antimicrobial compounds and an amino acid (serine) as most influential in predicting gestational age. Hypergeometric testing at the molecular family level additionally revealed that carnitine, bile acid, polyamine, nucleotide, penicillamine disulfide and chlorhexidine molecular families are most strongly associated with gestational age.

Although these metabolites may have a diverse range of biological functions and implications in a number of cellular processes, several have previously been reported to be implicated by the gut microbiome (amino and bile acids, carnitines, phosphatidylcholine derived compounds), involved in gut maturation (polyamines) or affecting gut microbial composition (antibiotics, diet-derived nucleotides) (40–43,47–51) (Figure 2, Supplementary Data 3, Supplementary Discussion).

In very recent studies, some of the microbiome-derived metabolites identified in our study have been shown to be associated with a diverse range of pathophysiologies related to preterm birth (Figure 2, Supplementary Data 3). There is a well-established increased risk for early- and also late-term complications to being born prematurely, so we can intuitively assume that risk biomarkers may be present in early life samples in prematurely born children. In our study, we identified increased relative amounts of several microbial catabolites in preterm children that have been linked to well-known late-term complications of prematurity, such as phenylacetylglutamine (associated with increased risk for cardiovascular disease (40,52,53)) or imidazole propionate (associated with type 2 diabetes (42,54)). Similarly, we found that kynurenic acid, a catabolite of tryptophan metabolism, is negatively correlated with gestational age, with higher relative abundances observed in preterm infants. Gut microbiota were shown to play an important role in tryptophan metabolism (41,47) and high levels of kynurenic acid in the central nervous system have been associated with schizophrenia and cognitive impairment (55). Although this was a methodological study and not designed to establish a direct causal or pathophysiological link between a specific marker and a later occurring complication, we may speculate that the observed difference in the neonatal metabolome may reflect an increased risk of early- or late-term complications on an individual level.

Common clinical practice such as antibiotics use seem furthermore reflected in the neonatal metabolome and significantly related to gestational age. Antibiotic-related metabolites (penicillamine disulfide and cefuroxime) and caffeine correlated negatively with gestational age, which reflects that preterm neonates are at increased risk of infection and often require broad-spectrum antibiotics from birth onwards (7). Caffeine is the most commonly used medication for treatment of apnea of prematurity (56). Antibiotics are known to have profound effects on gut microbial communities, and modified metabolic activity of the antibiotic-altered gut microbiome has been shown to result in decreased fecal levels of dipeptides and increased levels of primary bile acids and sugar alcohols in mice (51). In agreement with this finding, we here observed decreased blood levels of dipeptides and increased levels of primary bile acids and sugars, which could possibly result from an increased antibiotic use in prematurely born children. Similarly, gestational-age-dependent differential abundance of nucleotides among others may be reflective of differential feeding patterns. Preterm neonates are often fed parenterally, while term neonates are more likely to be breastfed.

Overall, we found more metabolites in preterm neonates when compared to term neonates. Within sample metabolite diversity on the other hand was not found to differ significantly across different categories of prematurity. Increased metabolite richness in preterm neonates could be reflective of increased medication in prematurely born children. Alternatively, it is known that intestinal permeability is higher in preterm compared to term neonates (57). Increased metabolite richness could thus also be reflective of increased intestinal permeability. The constant within sample metabolite diversity across different categories of prematurity would be in agreement with this hypothesis (more versus more diverse metabolites).

Our data reveals that metabolomic profiling of neonatal dried blood spots in combination with recently developed computational metabolome mining methods offers a powerful tool for monitoring gestational-age-dependent metabolic maturation in preterm neonates. Many of the metabolites here identified as significantly related to gestational age have previously been described to be related to the gut microbiome and short-as well as long-term health complications related to preterm birth. This finding is suggestive that catabolites of microbial metabolism are detectable in the neonatal blood as early as 2-3 days of life, and may play an important role in gestational-age-dependent metabolic maturation.

## Limitations

This study draws strength from being based on a large cohort of 148 samples, which currently is considered a large-scale study within the metabolomics field. Furthermore, the samples retrieved from the biobank were collected prospectively as part of the National Newborn Screening Program and therefore no systematic inclusion bias is likely to have influenced the study. Although we detected a large diversity of metabolites, metabolites extracted and detected in our study are inherently reflective of metabolites targeted during neonatal screening of inborn diseases. Similarly, only a limited number of metadata was available in this methodological study. We did not have access to data on future health outcomes, therefore our hypotheses regarding the role of the observed differences in metabolomics profiles of preterm versus term newborns are only speculative. A multi-omics approach and extensive prospective sampling would be needed to answer this question. Also, more samples would allow for more statistical power, whereas an independent test and training data set would allow for generalization of the findings outside of this study.

## Conclusions

Our data demonstrates that the neonatal metabolome is strongly reflective of gestational age. Many metabolites here found to be significantly associated with gestational age have previously been related to the gut microbiome or maturation and short-as well as long-term complications of preterm birth. This finding is suggestive that catabolites of microbial metabolism are detectable in the neonatal blood as early as 2-3 days of life, and may play an important role in gestational-age-dependent metabolic maturation. We show that metabolomic profiling of neonatal dried blood spots in combination with recently developed computational metabolome mining methods offers a powerful tool for monitoring metabolic maturation in preterm neonates. Further studies will be needed to establish direct causal or pathophysiological links between marker metabolites and later occurring disorders related to preterm birth.

## Data Availability

Data generated and/or analyzed in this study are not publicly available due to the risk of compromising individual privacy but are available from the corresponding author on reasonable request and provided that an appropriate collaboration agreement can be agreed upon.

## Acknowledgements

This research has been conducted using the Danish National Biobank resource, supported by the Novo Nordisk Foundation.

## Author contributions

ME performed statistical analysis and chemical structural annotation, interpreted the data and drafted the manuscript. SR performed statistical analysis, contributed to the interpretation of the data and the drafting of the manuscript. ULT contributed to data interpretation and the drafting of the manuscript. AB and SSL conceptualized and designed the study and acquired the data. JC contributed to data interpretation and the drafting of the manuscript. AB, MN, TW, PBM obtained funding. DH conceptualized and designed the study, obtained funding and together with ASC takes full responsibility for compliance with data sharing policies and ethical approval of the study. ASC conceptualized and designed the study and contributed to the interpretation of the data and drafting of the manuscript and together with DH takes full responsibility for compliance with data sharing policies and ethical approval of the study. All authors critically revised the manuscript for important intellectual content.

